# Drivers, Facilitators, and Impacts of Stigma Among Youth Living with HIV in Lima, Peru

**DOI:** 10.1101/2025.07.10.25331213

**Authors:** Marguerite Curtis, Alyson Nunez, Milagros Wong, Tiffany Chenneville, Kemesha Gabbidon, Trixie MacNeill, Renato A. Errea, Jerome T. Galea, Molly F. Franke, Kristin A. Kosyluk

**Author notes:** These authors have contributed equally to the work (Marguerite Curtis, Alyson Nunez).

## Abstract

Human Immunodeficiency Virus (HIV) stigma constitutes a major barrier to HIV prevention, testing, and treatment, and is associated with harmful consequences for people living with HIV such as discrimination, rejection from family and friends, and treatment nonadherence. This study aimed to understand the drivers, perpetuators, and outcomes of HIV-related stigma among young people living with HIV (YPLWH) in Lima, Peru. From November 2022 to June 2023, we conducted interviews and focus groups with a diverse group of 75 YPWLH, in addition to healthcare providers and HIV advocates, to discuss their experiences of HIV-related stigma. Audio recordings were transcribed verbatim and analyzed for emergent themes using framework analysis. Participants identified the drivers of HIV-related stigma as a lack of information about HIV and misinformation about HIV, and the key power groups who perpetuate this stigma, which included parents and older generations, educators, and healthcare providers. Participants reported outcomes of HIV-related stigma; these included feelings of fear, shame, and loneliness, and enacted behaviors such as treatment nonadherence, nondisclosure of HIV diagnosis, and social avoidance. Participants also reported health outcomes of sickness, death, and HIV transmission, and social outcomes including discrimination from employers, educators, and peers, and loss of social support. These findings are critical to understanding how HIV-related stigma manifests in the cultural context of Lima, Peru, and can be used to inform effective interventions to reduce and mitigate the impacts of HIV-related stigma.

## Introduction

Human Immunodeficiency Virus (HIV) stigma is a barrier to HIV prevention, testing, and treatment, resulting in preventable suffering and death [1]. In 2014, the Joint United Nations Programme on HIV/AIDS (UNAIDS) set the 95-95-95 targets, stating that 95% of people living with HIV (PLWH) should be diagnosed, 95% of those diagnosed should be receiving antiretroviral therapy (ART), and 95% of those receiving ART should achieve viral suppression by 2030 [2]. As national programs strive to close remaining service delivery gaps in pursuit of these targets, HIV-related stigma constitutes a major roadblock to these efforts and to the expansion of HIV prevention services, such as pre-exposure prophylaxis (PrEP) [3-4]. HIV-related stigma contributes to ART non-adherence, decisions about HIV disclosure to sexual partners, condomless sexual intercourse, loss of social support, and mental health problems [3], [5-10]. HIV-related stigma disproportionately impacts adolescents and young adults [11], a group already at high risk of unfavorable health outcomes [12-14]. Specifically, adolescents living with HIV (ALWH) can experience challenges transitioning to adult HIV care and have lower rates of ART adherence compared to adults [15-16]. For ALWH, stigma represents an additional burden during a period of pronounced physical, social, and psychological change [17]. HIV-related discrimination can result in loss of employment or housing, substandard medical care, and rejection by friends, family members and partners [18-19]. On the PLWH Stigma Index 2.0 [1], administered to 30,751 PLWH from 25 different countries, 85% of respondents endorsed some level of internalized HIV stigma, 13% reported experiencing discrimination from an HIV care provider due to their HIV status, and 24% reported experiencing HIV-related discrimination from some other type of care provider. Twenty-three percent of respondents reported discrimination in social settings, with rates of reported stigma and discrimination highest among transgender people and gay, bisexual men and other men who have sex with men (GBMSM).

Due to the social nature of stigma, a nuanced understanding of the ways that cultures stereotype HIV is required to effectively address HIV-related stigma and remove barriers to prevention and treatment [20-21]. One’s cultural identity contributes to experience of stigma [22]. In Latin America, HIV-related stigma is prevalent, with pervasive homophobia and exaggerated masculinity (i.e., machismo) exacerbating stigma towards the key populations of GBMSM and TGW [23-24]. In Peru, a 2023 survey found that half of PLWH reported discrimination related to their HIV diagnosis generally and 18% experienced discrimination from an HIV care provider. Feelings of self-stigma were reported by 76% of respondents and acts of self-exclusion by 33% [25]. Importantly, young people may experience HIV stigma in settings and through interactions distinct from those typically discussed in the adult literature [26]. For instance, stigma from peers, educators, and administrators in school environments is a significant concern for young people living with HIV (YPLWH) potentially affecting health, academic, and career outcomes [27]. In addition, transitions such as moving from pediatric to adult healthcare pose high risk periods where anticipated stigma may undermine treatment continuity.

The Health Stigma and Discrimination Framework is a global framework guiding how health-related stigmas manifest, their consequences, and potential interventions [28]. This framework takes a socioecological perspective to account for stigma’s complex drivers and facilitators, manifestations, and outcomes. To address a critical gap in the stigma literature, this study offers an innovative application of the Health Stigma and Discrimination Framework to examine how diverse groups of YPLWH in Peru experience HIV-related stigma. We aimed to 1) identify stigma’s drivers and facilitators (e.g., culturally bound stereotypes) in the Peruvian context from the perspective of YPLWH and various stakeholders, 2) identify stigma perpetuators (key groups that continue to perpetrate stigma), and 3) highlight key social and health outcomes impacted by stigma. By focusing on adolescents and emerging adults—a population often overlooked or insufficiently addressed in HIV stigma research—this study makes a significant contribution by advancing understanding of age-specific stigma dynamics within an under-researched regional context.

## Methods

### Study Setting and Population

Our work took place from November 2022 to June 2023 in Lima, where 40% of Peru’s estimated 110,000 PLWH reside [29]. The HIV prevalence in Peru’s general population is 0.4%, but is notably higher among transgender women (TGW, 32%), GBMSM (10%), and female sex workers (FSW, 2.4%) [29-30]. Around 57% of new HIV diagnoses annually occur in young adults ages 20-34 nationwide [31]. Venezuelan migrants to Peru living in Lima/Callao and Trujillo are particularly affected, with a prevalence of 1.01%, more than twice the national average among Peruvian adults (0.4%) and the adult population living in Venezuela (0.5%). For Venezuelan migrants, stigma due to HIV and migration status is a major barrier to care and treatment [32-33].

### Participants and Recruitment

We recruited a purposive sample of YPLWH ages 16-29 (n=75) through referrals from healthcare providers at public sector health facilities and contacts at community organizations working with key populations including GBMSM, TGW, FSW, as well as Venezuelan migrants, and cisgender females. While the majority of youth had recently acquired HIV, our sample also included a subset of young people who acquired HIV perinatally. In total, 34.7% of participants were assigned female at birth, 52% identified as gay or bisexual, and 12% identified as transgender (Table 1). Healthcare providers (n=11) and HIV advocates (n=24) ages 22-65 also participated.

**Table 1:** Participant baseline characteristics.

|  | YPLWH<br>(N=75) | Healthcare<br>Workers<br>(N=11) | HIV<br>Stakeholders<br>(N=24) | Total<br>(N=110) |
| --- | --- | --- | --- | --- |
| Characteristic | n (%) |  |  |  |
| *Sex assigned at birth* |  |  |  |  |
| Female | 26 (34.6) | 8 (72.7) | 5 (20.8) | 39 (35.5) |
| Male | 47 (62.6) | 3 (27.3) | 18 (75) | 68 (61.8) |
| Intersex | 2 (2.7) |  |  | 2 (1.8) |
| Other |  |  | 1 (4.2) | 1 (0.9) |
| *Gender identity* |  |  |  |  |
| Woman | 31 (41.3) |  | 6 (25) | 37 (33.6) |
| Man | 33 (44) |  | 10 (41.7) | 43 (39.1) |
| Transgender woman | 9 (12) |  | 6 (25) | 15 (13.6) |
| Genderfluid | 1 (1.3) |  | 1 (4.2) | 2 (1.8) |
| Gender nonbinary |  |  | 1 (4.2) | 1 (0.9) |
| Other category | 1 (1.3) |  |  | 1 (0.9) |
| Data not available |  | 11 (100) |  | 11 (10.0) |
| *Ages* |  |  |  |  |
| 16 - 18 | 8 (10.7) |  |  | 8 (7.3) |
| 19 - 29 | 66 (88) | 1 (9.1) | 6 (25) | 73 (66.4) |
| 29 < | 1 (1.3) | 9 (81.8) | 18 (75) | 28 (25.5) |
| Data not available |  | 1 (9.1) |  | 1 (.9) |

### Data Collection

A Peruvian certified nurse (MW) experienced in conducting qualitative research with YPLWH conducted 13 focus groups and 16 individual interviews. A semi-structured interview guide based on the Health Stigma and Discrimination Framework [28] was used to explore participant experiences with stigma, manifestations of stigma in Peru, and the perceived etiology of stigma (Supplementary Material). Prior to data collection, MW received training on the framework components and their application to the guide. Ten focus group sessions were held in person at Socios En Salud (Partners In Health) offices in Lima. Three focus groups and all interviews were conducted virtually. Sessions lasted approximately 90 minutes and were audio recorded. No notes were taken. During five larger in-person focus group sessions, an additional team member was present to facilitate. There were no follow-up sessions. Transcripts were translated verbatim from Spanish to English and were not returned to participants for review. Participants were reimbursed for transportation costs and received 50 soles (13 USD) for their time.

### Data Analysis and Reporting

We adhered to COREQ guidelines for the reporting of qualitative research [34]. Audio recordings were stored on a password-protected encrypted server only accessible to study team members and transcripts were uploaded into Dedoose for analysis [35]. Data were analyzed and grouped into emergent themes using Framework Analysis [36-37] guided by the Health Stigma and Discrimination Framework. MW and project team member (AN) coded the data in Spanish using a preliminary codebook derived from the focus group/interview guide, adding *de novo* codes as needed. Both coders reviewed and coded each transcript independently, and then discussed code application and any discrepancies to reach consensus on all code applications. Our approach prioritized researcher reflexivity and collaborative analysis over strict computing of interrater reliability scores.

### Ethical Considerations

The Vía Libre Bioethics Committee in Peru and Harvard Medical School Institutional Review Board approved this research. Participants provided informed consent and caregiver consent with participant assent was obtained for those <18 years of age. We obtained a waiver of consent for minor YPLWH who had not disclosed their diagnosis to guardians. The data are not publicly available due to privacy restrictions.

## Results

The Health Stigma and Discrimination Framework describes how drivers and facilitators of stigma lead to stigma marking (the process of applying stigma to people or groups), stigma experiences and practices, and subsequent outcomes for affected populations, organizations and institutions. Here we present the principal drivers (lack of information and misinformation) and perpetuators (older generations, educators, healthcare providers, and power groups) of HIV-related stigma in Lima, Peru and the health and social outcomes this stigma has for YPLWH. Additional illustrative quotes are provided in Supplementary Table 1.

### Drivers of stigma

Drivers of stigma are factors that facilitate health-related stigma or mediate stigma marking. Reported stigma experiences were associated with two primary drivers: a total lack of information about HIV and misinformation about HIV. These drivers emerged among family members, healthcare providers, educators, other power groups, and YPLWH themselves.

#### Lack of information about HIV

Participants cited a lack of information regarding transmission, treatment, and the lived experience of PLWH as a principal driver of stigma. They mentioned that societal taboos can preclude open discussion of HIV and sex in educational or familial settings, leading to a general lack of awareness that is transferred across generations. Participants also emphasized that living with HIV is not synonymous with being well-informed about HIV. They perceived that the healthcare system provides insufficient education to individuals newly diagnosed with HIV, leaving them to understand their condition however they can; others may choose not to inform themselves. Furthermore, participants noted that those who do not perceive themselves as at risk for HIV may feel the topic is beneath them and lack motivation to educate themselves, further perpetuating stigma:

> *The community has the wrong idea about the condition and how it’s transmitted, and that’s because the education system doesn’t teach that information in schools. It’s like it’s a taboo that you can’t talk about. (YPLWH, cisgender woman)*

> *Having the diagnosis doesn’t necessarily mean you’re informed, I think each of us has learned about it as best we could. We haven’t always had someone to give us a presentation or a class at university or school or even our parents. A lot of times we’ve had no one and we’ve come across a video on YouTube, or maybe we have searched for our own information. And there are other people who have no intention of informing themselves. So being seropositive isn’t a synonym of, isn’t a guarantee that someone knows about the topic. (YPLWH, GBMSM)*

#### Misinformation about HIV

Misinformation as a driver of HIV-related stigma includes erroneous and/or outdated beliefs or stereotypes perpetuated by influential entities or actors, such as educators, healthcare providers, and other power groups. Participants explained that stigma in Lima stems from stereotypes about modes of transmission, what living with HIV looks like, and the relationship between HIV and the LGBTQIA+ community. They described the general population as believing that HIV can be transmitted through physical proximity to PLWH and placed responsibility on the education system for not providing HIV-related instruction, particularly regarding the topics of transmission and detectability:

> *And if someone says, be scared of this person because they have something that can kill you, people think ‘he’s going to kill me, he’s going to give it to me, I’m going to get it’. So, in their ignorance, they think it’s dangerous. But since they haven’t researched, they don’t know, they can’t be sure if what they’re being told is true or not. (YPLWH, GBMSM)*

The media, specifically movies depicting PLWH, fails to reflect recent medical advances and chooses to portray PLWH as sick, frail, and on the verge of death. Participants believe this contributes to pervasive beliefs that PLWH cannot lead productive lives or have partners or children. One participating healthcare provider mentioned that parents learning of a child’s HIV diagnosis sometimes even ask themselves “what’s the point of sending my child to school?”, and as one HIV advocate notes:

> *They think because someone’s chubby ‘oh no, he can’t have it [HIV] because he’s chubby, full of life’. We have this prejudice many times. And when they see someone who’s thin ‘he probably has HIV, I’m going to use a condom’. I mean that comes from the media, how they’ve portrayed this condition, that’s why many people have these ideas. (HIV advocate)*

According to participants, the erroneous belief that HIV only affects the LGBTQIA+ community is maintained by educators and family members who believe outdated information. Participants explained that this is correlated with judgments about PLWH’s life decisions and morality. Participants mentioned the intersection of education and religion, stating that religion portrays HIV as a punishment from God and PLWH as deserving of the condition:

> *[It comes from] an educational construct from childhood, when they give those HIV talks at school saying that it’s a disease that’s usually transmitted in the gay community. And from the religious angle … it’s a punishment from God. I’ve heard that many times. (HIV advocate)*

### Key power groups perpetuating stigma

Facilitators of stigma include individuals or groups who have the power—referred to as “stigma power” — to “mark” or apply stigma to PLWH [38-39]. According to our participants, facilitators perpetuate stigma driven by a lack of information and beliefs in outdated or erroneous information. Participants identified parents/family members, older people, educators, healthcare providers, and other power groups (religion, employers, the media, and government) as groups that perpetuate stigma in Lima. Stigma marking occurs from direct interactions with PLWH or as a result of social and cultural norms or health and employment policy, which results in stigma being applied to PLWH (Figure 1). The misinformation is perpetuated across generations, which facilitates the continuation of stigma and its deleterious consequences for PLWH and HIV prevention efforts. Inaccurate information is the driver of stigma. The perpetuation of that inaccurate information represents a potential moderator. Key power groups are possible sources of moderation because the community looks to these key groups for information when formulating their own opinions about social issues, such as HIV. Therefore, people in positions of power within a community are positioned to moderate levels of HIV-related stigma by either perpetuating erroneous and outdated information or rectifying stigmatizing beliefs and attitudes.

**Figure 1.**
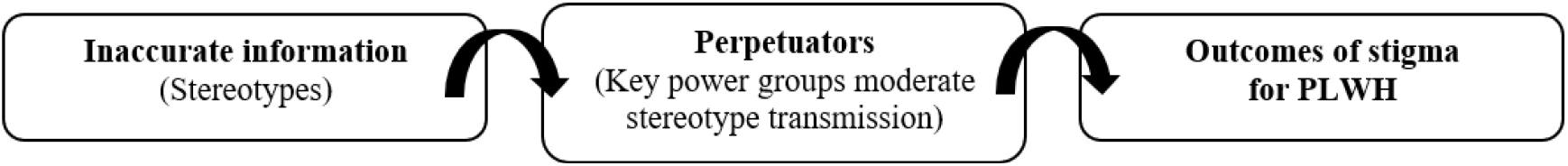
Moderation of stigma.

#### Parents & older generations

Participants identified parents and older people generally as sources of stigma because of their tendency to maintain outdated beliefs regarding HIV. They perceived these generations as less able or unwilling to assimilate current information about HIV into their way of thinking, resulting in stigmatization. Some participants attributed these patterns to conservative religious identities or to the machismo pervasive in Peruvian culture, while others highlighted older people’s lack of engagement with information available on new technological platforms. Accordingly, participants described feeling more comfortable interacting with younger healthcare providers as they tend to be more knowledgeable about HIV medical advances and more empathetic:

> *Adults in general are a little more closed-minded to understand things, whereas young people are already familiar with certain information. That’s in general, not just doctors but really any profession. It’s harder for adults to process this type of information. (HIV advocate)*

> *I’ve experienced quite a lot of discrimination from doctors and in the clinic. I don’t know why, because they’ve studied… The younger doctors I do appreciate, because my doctor always tells me … “keep your head up, you’re the same as me, there’s no reason to think of yourself as less than*.*” (YPLWH, cisgender woman)*

#### Educators

Participants described educational institutions as either failing to educate on HIV or perpetuating HIV misinformation. In institutions where HIV was discussed, participants noted educators were themselves uneducated about HIV and, therefore, perpetuated harmful stereotypes in their classrooms. They described this as being particularly dangerous as students tend not to question information presented by a teacher, especially at the primary and secondary levels, reinforcing power dynamics and how they contribute to HIV-related stigma. Educators facilitate stigma marking when they present misinformation, fail to correct erroneous or outdated information, and fail to support students who reported feeling isolated or discriminated against:

> *I studied at a public school and nothing, absolutely nothing was said [about HIV]. (YPLWH, cisgender woman)*

> *At school, when we were in the HIV/AIDS talk, the teacher didn’t distinguish between HIV and AIDS because she just said, ‘people who have it, have AIDS*.*’ And I sat there thinking and I said no. I raised my hand and said, ‘no because AIDS is at the end and HIV is when you have treatment*.*’ And my friend said, ‘no miss, … I’ve researched the topic and HIV has treatment but it doesn’t have a cure. If you don’t take your medicine every day, there’s a chance you’ll go into the final stage which is AIDS*.*’ And the teacher didn’t know that. So. I mean, seeing as she’s a teacher, she should research a little. (YPLWH, perinatal transmission)*

#### Healthcare providers

Participants considered healthcare providers to be an influential group perpetuating stigma. They explained that though providers tend to be more informed about HIV than the general population, this information is not necessarily enough to counter personal prejudices. Additionally, they mentioned that not all healthcare personnel interfacing with PLWH at hospitals and clinics have received appropriate training, with some manuals used continuing to present outdated information. One participant noted that she has had to explain the term undetectable to multiple non-specialist doctors. Many participants also described discrimination faced in terms of questions, comments, attitudes, and body language towards PLWH during medical visits, as some healthcare personnel believe that PLWH are deserving of their diagnosis and undeserving of HIV care. Healthcare providers themselves mentioned a lack of doctors educated on the specific needs of GBMSM and TGW and that some colleagues see working with PLWH as a punishment:

> *It’s true that healthcare personnel have a certain level of information, but information doesn’t uproot personal prejudices. (Healthcare provider)*

> *There’s a group of doctors who still have the idea that HIV is a highly lethal, highly contagious virus, as if we were in the 90s. And they still believe that now. They think it shouldn’t be a priority of the public healthcare system to treat [PLWH], they think [PLWH] should be responsible for their own medical expenses and should find treatment however they can. So it’s that erroneous sharing of information, treating it as the truth and not taking the time or the bother to research a bit more or to separate their ideological or political bias. (YPLWH, GBMSM)*

> *I was in the operating room and two nurses came in to help the doctor. And you know what one of them said?*

> *‘Ah, she has HIV, don’t touch her*.*’ (YPLWH, TGW)*

#### Other key power groups (religion, government, the media, employers)

Other power groups were identified as upholding social, gender, and cultural norms or perceived occupational safety standards that perpetuate HIV-related stigma. According to participants, religious groups viewed homosexuality as a sin and HIV as the consequence of this sin and disseminated this message to their congregations. Participants believe government agencies and civil society take little interest in HIV-related issues and, therefore, perpetuate stigma by failing to act on behalf of PLWH. The media presents outdated information and images of what living with HIV looks like, contributing to stigmatizing behaviors and attitudes on the part of the uninformed population:

> *So then you have the religious people who say ‘oh they’re devil-worshippers, that’s why they’re like this’ … and ‘they’re sick and they deserve it, that’s their punishment*.*’ …And it leads to families saying ‘I hope this never happens to us’ and then it happens. They have a child with HIV, and with the diagnosis and their sexual orientation, they’re discriminated against [by their families]. (YPLWH, GBMSM)*

> *There’s also a lot of incorrect information on the Internet and on TV being shown to people. And there are people that that’s what they believe. (YPLWH, perinatal transmission)*

Additionally, among these other power groups, participants illustrated the institutional policies that facilitate stigma. For example, many participants identified discriminatory attitudes and prejudices from their employers. However, some participants who referenced their experiences disclosing their diagnosis in the workplace did not speak to explicit acts of discrimination from their employers. Instead, institutional factors, like requiring time off to pick up ART and needing to disclose reasons for this time off, perpetuated the stigma:

> *The issue of asking for time off at work, especially being a migrant, knowing that this opens the door to you getting fired, having to give explanations, having to get permissions, saying you’re sick when really you’re just going to pick up your treatment. It’s complicated. (HIV advocate)*

### Outcomes of Stigma Among PLWH

Among participating PLWH, we found a principal outcome of HIV-related stigma to be the internalization of stigma; this internalized stigma creates feelings of fear, anxiety, shame/guilt and loneliness. These feelings drive certain behaviors among PLWH, including choosing not to seek out or adhere to treatment, nondisclosure of HIV diagnosis, and self-isolation. Together with stigmatizing and discriminatory treatment from family, friends and healthcare providers, these behaviors contribute to a variety of health and social impacts for PLWH including sickness, death, loss of educational and employment opportunities, and damaged relationships with friends, family and sexual partners. Here, we demonstrate the process by which the feelings caused by stigma and discrimination drive behaviors for PLWH, both of which produce health and social outcomes (Figure 2).

**Figure 2.**
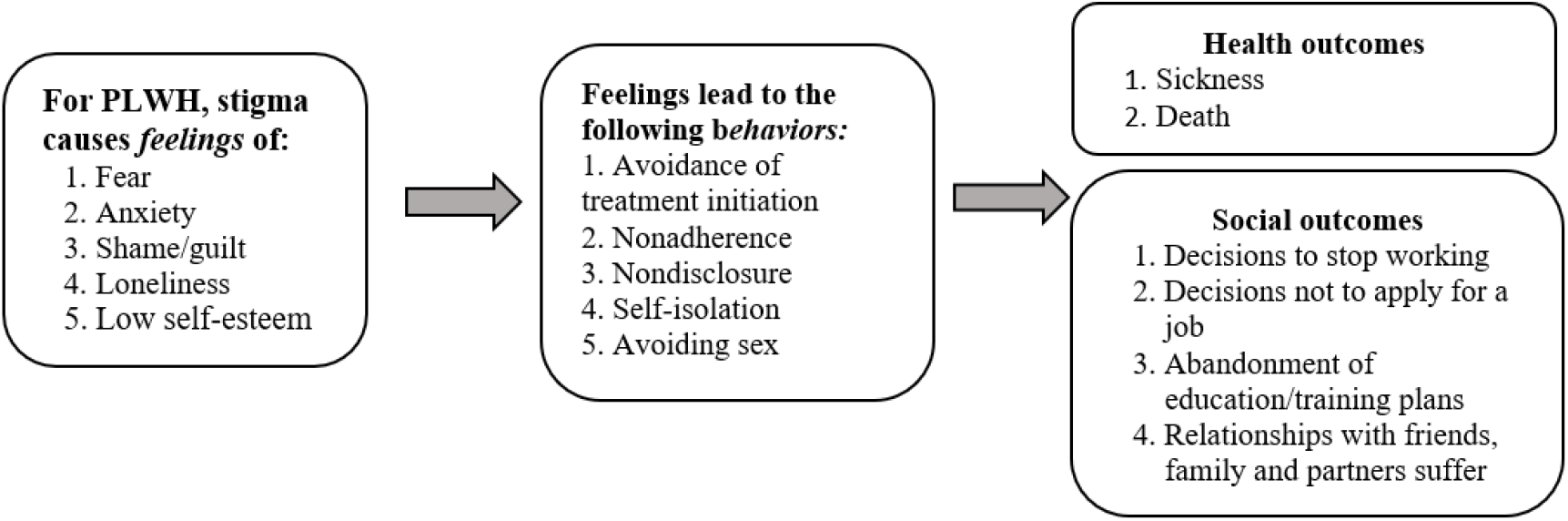
Outcomes of Stigma among PLWH.

#### Feelings of fear and anxiety

Fear was attributed to misinformation surrounding the diagnosis; for example, participants believed that receiving an HIV diagnosis was a death sentence or would lead to severe illness or feared transmitting HIV to others in ways that science does not support (e.g., sharing utensils). Participants described fear of their diagnosis being discovered and of discrimination due to their diagnosis, including being treated as immoral or contagious. As a result of this fear, participants discussed anxiety related to hiding their HIV diagnosis, and the anticipation of discriminatory or stigmatizing behavior. Finally, participants described anxiety resulting from fear of transmission, even if this fear (e.g. a fear of transmitting HIV through accidentally cutting oneself) was rooted in misinformation. These feelings, often informed by past experiences, were particularly notable in professional and healthcare settings:

> *You don’t know how other people are going to react when they find out about something, but there’s still that discrimination, that fear, and it does affect you … it’s not directly that they’ve discriminated against you, but for example it’s what they might do if they found out. (YPLWH, perinatal transmission)*

> *For young people, it’s complicated, especially when you’re a minor. Because for example in my case, it was always thinking about [HIV], associating it with death and thinking that I wouldn’t make it to 40 and that my studies would be cut off, that I wouldn’t be able to study, I wouldn’t be able to have partners, which are the constant fears that appear when you’re very young because you don’t have a lot of information. (HIV advocate)*

> *When I was cutting [my daughter’s] nails and I cut myself, oh my God, oh my God. Maybe it’s hard to explain but the fear [of transmitting HIV], the suffering, it’s awful, awful. (YPLWH, FSW)*

#### Feelings of shame, guilt, and loneliness

Participants explained how HIV-related stigma created feelings of shame, guilt, or self-disgust primarily due to others treating PLWH as infectious or contagious. For example, participants discussed how shame was brought on by others’ distancing and avoiding contact —hugging, kissing, or sharing food— with PLWH, rooted in misconceptions about HIV transmission. Additionally, PLWH, regardless of being undetectable or taking precautions to prevent transmission, dealt with a lack of trust or direct blame from others, with one participant noting that stigma creates a situation of “constant shame” for PLWH while also increasing their own anxiety around transmission (HIV advocate). This mistrust and assigning of blame by friends, family members, partners, healthcare providers, and even to oneself, speaks to the social rejection and exclusion PLWH experience because of their diagnosis. Consequently, participants described suffering from loneliness, and often having to navigate treatment alone, even at a young age:

> *A friend I told did distance herself from me, she doesn’t talk to me anymore. She thinks that if we drink out of the same glass, she’s going to get [HIV], or if I take some food from her, she’s going to get it. That part did affect me, and you feel embarrassed too because it’s out of nowhere. We used to share everything, … She wears a mask now. (YPLWH, cisgender woman)*

> *They put us in a situation of constant guilt, which doesn’t allow us to enjoy even our sexuality, really anything. For example, when I came out the first thing people asked was if I was going to get HIV and this was probably because my family had had some experiences related to that, beliefs, erroneous beliefs… This creates trauma in our sexuality which weighs on every sexual relationship all the time. (HIV advocate)*

#### Behaviors

Participants described how HIV-related stigma drove certain behaviors, including avoidance of HIV testing and treatment initiation, HIV treatment nonadherence, nondisclosure of HIV diagnosis, and self-isolation, including the avoidance of sexual encounters. Multiple participants discussed how stigma discouraged some young people from getting tested, with one activist pointing out that many people “would rather die not knowing,” keenly aware of the burden of a highly stigmatized HIV diagnosis. PLWH and HIV providers noted that not initiating or adhering to treatment was often motivated by fear of being recognized at an HIV treatment facility and fear of discrimination by healthcare professionals. In addition to treatment avoidance, many PLWH discussed fear of disclosure due to stigma and anticipation of stigmatizing behavior, such as assumptions about how HIV was acquired. Some participating PLWH felt the need to hide or socially isolate themselves in order to avoid being a target of stigma. HIV-related stigma also had the potential to impact sexual behavior, as participants reported avoiding sex for fear of transmitting HIV to sexual partners:

> *I wasn’t getting treatment precisely because of that fear that someone would see me, someone would recognize me. And a few times I would get to a [health center] and I wanted to start treatment and ‘oh, I know him, I have to leave*.*’ (YPLWH, GBMSM)*

> *Discriminating against patients at health centers, what it does is it pushes them away. It pushes them away from services and these patients leave with more questions than they came with many times. So, they don’t get testing, they don’t get treatment, they don’t get counseling, they don’t get prevention methods. (Healthcare provider)*

> *And so from that moment [of the HIV diagnosis], I’ve isolated myself from everyone. I haven’t wanted to have a partner or anything because I feel a responsibility that if I’m going to fall in love or be with someone, that responsibility that at any moment I could give it to them or I could ruin their life and I don’t want that. I still feel that way today. (YPLWH, cisgender woman)*

#### Health Outcomes

Negative feelings and the associated behaviors resulting from HIV-related stigma have an important impact on health outcomes for PLWH and their sexual partners, including sickness and death. Participants explained how stigma and the discriminatory treatment of PLWH in healthcare settings negatively impact treatment adherence and prevent individuals from getting the care they need. Sometimes, HIV-related stigma impacts access to other healthcare services. For example, participants described being denied care, charged an extra fee due to their HIV diagnosis, or being told an HIV diagnosis would prevent them from accessing other routine healthcare services such as dental care:

> *[The doctor] came to my house and knocked on the door and said, ‘yes, you’re positive. Take this and don’t come back to the clinic because people sick like you can’t come to the clinic since you’re contagious, and much less have appointments in the dental office*.*’ She said, ‘you don’t belong to the clinic anymore… you have to go to a health center in Villa El Salvador called San José. Go there and tell them what you have, that you’re positive, and there you’ll get your treatment. Don’t ever come back to the clinic*.*’ (YPLWH, FSW)*

Participants also mentioned the negative impacts of stigma and associated behaviors on mental health, including depression, anxiety, and suicidal ideation. They shared experiences with suicidal thoughts and attempts due to HIV-related stigma and experiences of PLWH in their life who died by suicide or AIDS-related causes due to treatment nonadherence stemming from mental health conditions:

> *When I got my diagnosis, my world came crumbling down. I was alone, I only had my roommate. And one day my roommate found me when I had overdosed on pills because I wanted to kill myself, I didn’t want to go on. I mean I’m really young and I didn’t know how I was going to tell my family, ‘look, this is what’s happened*.*’ It wasn’t in my plans. (YPLWH, FSW)*

> *[My best friend] starts to lose weight, he was thinner, he got sick from anything. And I asked myself, ‘what’s happening to him?’ And that’s when he tells me he has HIV and that he’s not taking the treatment. And I ask him why. Physically and emotionally, mentally, he was really bad, really really bad. He didn’t have the support of his family, he didn’t have any money, and that ended up affecting him to the point that he was hospitalized. I don’t remember if it was 2018, but it wasn’t because of HIV specifically, I don’t remember the disease, I think maybe tuberculosis. But literally that’s why he died. His family never supported him, his friends left him completely alone, no one wanted to take care of him. (HIV advocate)*

#### Social Outcomes

In addition to health outcomes, participants discussed the social outcomes of HIV-related stigma, which significantly impacted how PLWH work, study, and maintain relationships with family, friends and partners. HIV-related stigma presents challenges for PLWH in the workplace. For example, participants described how having an undisclosed HIV diagnosis can be a major source of anxiety for PLWH who might be required to leave work periodically for medical appointments. Additionally, some workplaces maintain outdated and stigmatizing policies that require employees to undergo unnecessary HIV testing, challenging disclosure autonomy and placing them at risk for discriminatory action. Many participants cited fear of requesting permission to leave work for medical appointments and denial of promotions or career advancement opportunities. For many participants, this stigma, or the anticipation of stigma, significantly impacted their decisions surrounding work, with some opting not to apply for a job that they desired. Similarly, participants also shared experiences of stigma by educators and peers in the school setting, sometimes pushing PLWH to abandon education:

> *[Potential employers] trick you by saying that they’re not going to ask for it [an HIV test] but in the end they do and that’s why you hold yourself back, you tell yourself you can’t do it. I’ve studied to be a pastry chef… but unfortunately I can’t work. (YPLWH, cisgender woman)*

> *I had invited [classmates] to my house to do group work since we could use the internet and they agreed. Two ended up coming and they started going through my things, moving them, and when they’re moving them they find my medicine since I usually put it next to my papers. And one of them takes it …and says, ‘you have HIV? Are you crazy? And I say, ‘Wait what? No, I don’t understand*.*’ I didn’t want to say yes or anything. I said, ‘I don’t understand you*.*’ And she took the paper, showed it to me and said, ‘you disgust me*.*’ She wrinkled it up and threw it at me and left with the other girl. And then they went and told all of my classmates that I had HIV and they all started, ‘no don’t come near me*.*’ (YPLWH, FSW)*

Participants noted how HIV-related stigma also had a profound impact on their social relationships. Upon receiving a diagnosis, many participants were stigmatized and rejected by family, friends and partners. Many participants reported distancing by family and friends following disclosure of their HIV diagnosis; often this distancing coincided with participants’ own avoidance of these relationships due to depression, shame, and fear of stigma:

> *After that [my diagnosis], I’ll be honest, my world came crumbling down. I was prescribed a medicine to take, three pills, and I have to take it every night. And I try to deal with it. I haven’t had a romantic relationship with anyone, I have to keep working even though I don’t want to. I have to keep doing it because I have to study. I have my mom who’s upset, my sister that doesn’t want to talk to me, but she has three kids and I support her and she doesn’t even want to see me. So, I have to keep going and it’s very, very difficult. And there are moments when I just want to stop taking my medicine, I want to stop and give up. And then I say no. (YPLWH, FSW)*

> *I didn’t like going to parties, I didn’t like going to school events, I didn’t have friends. I shut myself up at home with my sister and she was the one who defended me, but I didn’t make friends. I became very antisocial. I think I grew up very antisocial because anyone who got close, they already knew because my parents died [of HIV]… It affected me a lot emotionally that I didn’t want to have friends, I didn’t want to go out. I became very bitter you could say. (YPLWH, perinatal transmission)*

## Discussion

Consistent with research in other resource-constrained settings (e.g., Kenya ([40] and India [41]), our participants identified several key drivers of stigma, including misinformation about routes of transmission, and stereotypes that HIV is a fatal condition, that PLWH appear sick, that HIV is a condition only affecting the LGBTQIA+ community, and that PLWH are deserving of the diagnosis due to immoral or promiscuous behavior. Tattsbridge et al [42] also identified misinformation and misperceptions about HIV as key drivers of stigma among MSM in Loreto, Peru, suggesting the need for education campaigns across groups and regions in Peru.

Key power groups potentially moderating community levels of stigma by facilitating the perpetuation of stereotypes and misinformation included parents and older generations, educators, healthcare providers, religious institutions and leaders, employers, landlords, the media, and government officials. Valencia-Garcia et al [43] also identified healthcare providers as a key power group that perpetuates the stigma surrounding HIV based on interviews with women living with HIV in Lima. The central role of healthcare providers, families, and communities in perpetuating HIV stigma through negative labeling, avoidance, and denial of treatment, often driven by misinformation, fear, and sociocultural norms is documented in other settings as well, highlighting the potential for these key groups to minimize stigma through education, awareness, and professional support [44].

Outcomes of HIV stigma included the internalization of stigma, resulting in feelings of fear, anxiety, shame, and loneliness. These feelings lead to behaviors that negatively impact outcomes for YPLWH including choosing not to seek treatment, treatment nonadherence, nondisclosure of HIV status, self-isolation, and avoiding sex [45-46]. Key outcomes of stigma for YPLWH identified by our participants include health (i.e., physical and mental health) and social outcomes (i.e., work and education). These findings align with previous literature suggesting poorer physical health, more depression symptoms, higher emotional distress, and poorer overall quality of life resulting from HIV-related stigma [47]. Further, these findings also support prior research documenting the experiences of other demographic groups of PLWH in Peru. Based on interview data from MSM living in Loreto, Peru, Tattsbridge et al [42] uncovered a culture of isolation and stigma, resulting in shame surrounding HIV status and impact on employment. Valencia-Garcia et al [43] documented maltreatment and violation of rights and privacy as an outcome of healthcare provider stigma among women living with HIV in Lima. Combined with existing research, current findings suggest that the negative outcomes of HIV stigma in Peru are widespread among PLWH regardless of age, gender, and sexual orientation. The use of the Health Stigma and Discrimination Framework to guide data collection and analysis permitted a greater understanding of the relationship between drivers and facilitators of stigma and outcomes for YPLWH and the key power groups involved in the HIV stigma process.

Understanding how HIV stigma manifests in the cultural context of Lima, Peru, from the perspective of YPLWH is key to developing interventions that will be effective in reducing local HIV stigma and preventing negative health and social outcomes. Interventions must be designed to tackle the predominant stereotypes driving the stigma process and pay special attention to key power groups that perpetuate stereotypes and shape community perceptions. Interventions should reflect a consideration for YPLWH’s critical developmental stage along with existing socio-cultural inequities that produce unique or heightened effects of stigma [48]. Interventions should be designed with outcomes in mind [49]. Our findings point to key health and social outcomes that should be considered as metrics of success of interventions. Specifically, successful interventions should reduce internalized stigma among PLWH, resulting in improvements in self-esteem, and reductions in fear, anxiety, shame, loneliness, and sadness [50]. Other stigma work addressing internalized stigma has focused on measuring the stress associated with these feelings— stigma stress—as a key outcome of internalized stigma reduction efforts [51]. Behavioral outcomes resulting from successful interventions should include increased treatment seeking and adherence, social inclusion, and safe and healthy sex practices. Ultimately, interventions should bring about improvements in physical and mental health, reductions in death by AIDS and suicide, and reduced HIV transmission, as well as improvements in social outcomes such as employment, training and education, and relationships. Improved quality of life among YPLWH also is a laudable goal of HIV stigma-reduction interventions [52].

According to the Health Stigma and Discrimination Framework, “facilitators” of stigma are more than just key power groups, and include structural facilitators such as cultural norms, social and gender norms and equality, occupational safety standards, legal environment, and health policy [28]. Policy work to understand how occupational standards and health policies might be perpetuating stigma and how stakeholders might work to affect policy change to remediate the adverse effects of structural stigma are important areas for future work. A limitation of this research is that while our interview guide evoked participant responses that elicited the identification of key power groups, we could not draw conclusions about other structural facilitators. Participants may have been unaware of the structural forces at play, or there may be alternative questions that need to be asked in order to elicit participants’ understanding and experiences with structural stigma. Future research should delve deeper by asking questions to identify these other facilitators.

Another limitation of the current study is that data do not allow for an examination of intersectional experiences of stigma in a substantive way. The duration of our focus group and interview sessions did not allow for us to probe about the cumulative effect of stigma associated with multiple marginalized identities. Future work should aim to capture intersectionality in order to understand how it impacts stigma experiences among YPLWH and how it might best be addressed. An additional limitation of this work is that our Lima sample may not be representative of other areas of Peru. Stigma seems to be even greater in the tropical and mountain regions outside of Lima according to our participants, who mention that these populations tend to be generally more conservative.

In conclusion, we found that HIV stigma among YPLWH in Lima, Peru is driven by stereotypes that HIV is a fatal condition or that PLWH appear sick, misinformation about transmission, and moralistic narratives that blame individuals for their diagnosis. These misconceptions are perpetuated by key power groups, leading to internalized stigma, fear, anxiety, shame, loneliness, and reduced self-esteem. As a result, YPLWH may avoid treatment, struggle with medication adherence, refrain from disclosing their status, self-isolate, or avoid sex, contributing to both negative health outcomes and social consequences [47], [53-54]. Designing stigma interventions with an understanding of the cultural context and how stigma manifests locally promises the greatest improvement in outcomes [49]. These findings will facilitate the design of targeted multisectoral and multi-level HIV stigma interventions in Peru and elsewhere in Latin America.

## ETHICS APPROVAL

Approval was obtained from the Peruvian Vía Libre Bioethics Committee and from the Harvard Medical School Institutional Review Board in September 2022. The procedures used in this study adhere to the tenets of the Declaration of Helsinki.

## CONSENT

Participants and guardians of participants <18 years old provided written informed consent or assent, as applicable. The study was granted a waiver of consent for adolescents <18 years old who had not disclosed their HIV status to a guardian.

## DATA AVAILABILITY

The data that support the findings of this study are available upon reasonable request from the corresponding author. The data are not publicly available due to privacy or ethical restrictions.

## AUTHORS’ CONTRIBUTIONS

Study design: M.W., K.K., J.G., M.F., R.E.

Analysis design: K.K., J.G., M.F., R.E.

Study implementation: M.W.

Data management: A.N., M.C., M.W.

Data analyses: A.N., M.C., T.M., M.W., K.K., R.E.

Data interpretation: all authors

First draft: A.N., M.C., T.M., K.K, M.F., R.E.

Critical review of manuscript: all authors

Approved final version: all authors

## ACKNOWLEDGEMENTS

The authors are indebted to the study participants, and to the healthcare providers and community partners who supported the study team in participant recruitment. We would also like to express our gratitude to the Fogarty International Center for their financial support and to the Socios en Salud Youth Advisory Board for their guidance throughout our project.

## References

[1] Global Network of People Living with HIV (GNP+), “People Living with HIV Stigma Index 2.0. Global Report 2023. Hear Us Out: Community Measuring HIV-Related Stigma and Discrimination.,” Amsterdam, 2023.

[2] UNAIDS, “Understanding Fast-Track: accelerating action to end the AIDS epidemic by 2030.,” 2015.

[3] S. M. Sweeney and P. A. Vanable, “The Association of HIV-Related Stigma to HIV Medication Adherence: A Systematic Review and Synthesis of the Literature.,” AIDS Behav, vol. 20, no. 1, pp. 29–50, Jan. 2016, doi: 10.1007/s10461-015-1164-1.

[4] A. G. Perez-Brumer et al., “Homophobia and heteronormativity as dimensions of stigma that influence sexual risk behaviors among men who have sex with men (MSM) and women (MSMW) in Lima, Peru: a mixed-methods analysis.,” BMC Public Health, vol. 19, no. 1, p. 617, May 2019, doi: 10.1186/s12889-019-6956-1.

[5] N. Ammon, S. Mason, and J. M. Corkery, “Factors impacting antiretroviral therapy adherence among human immunodeficiency virus–positive adolescents in Sub-Saharan Africa: a systematic review,” Public Health, vol. 157, pp. 20–31, Apr. 2018, doi: 10.1016/j.puhe.2017.12.010.

[6] M. S. McHenry et al., “HIV Stigma: Perspectives from Kenyan Child Caregivers and Adolescents Living with HIV,” Journal of the International Association of Providers of AIDS Care (JIAPAC), vol. 16, no. 3, pp. 215–225, May 2017, doi: 10.1177/2325957416668995.

[7] C. Haines, M. E. Loades, B. J. Coetzee, and N. Higson-Sweeney, “Which HIV-infected youth are at risk of developing depression and what treatments help? A systematic review focusing on Southern Africa,” Int J Adolesc Med Health, vol. 33, no. 5, Oct. 2021, doi: 10.1515/ijamh-2019-0037.

[8] J. T. Galea et al., “Barriers and facilitators to antiretroviral therapy adherence among Peruvian adolescents living with HIV: A qualitative study,” PLoS One, vol. 13, no. 2, p. e0192791, Feb. 2018, doi: 10.1371/journal.pone.0192791.

[9] C. A. Rodriguez et al., “Understanding health-related behavior among adolescents living with HIV in Lima, Peru,” BMC Pediatr, vol. 19, no. 1, p. 396, Dec. 2019, doi: 10.1186/s12887-019-1773-3.

[10] C. F. Cáceres, P. Aggleton, and J. T. Galea, “Sexual diversity, social inclusion and HIV/AIDS,” AIDS, vol. 22, no. Suppl 2, pp. S45–S55, Aug. 2008, doi: 10.1097/01.aids.0000327436.36161.80.

[11] D. Kerrigan, A. Vazzano, N. Bertoni, M. Malta, and F. I. Bastos, “Stigma, discrimination and HIV outcomes among people living with HIV in Rio de Janeiro, Brazil: The intersection of multiple social inequalities,” Glob Public Health, vol. 12, no. 2, pp. 185–199, Feb. 2017, doi: 10.1080/17441692.2015.1064459.

[12] R. S. Boerma et al., “Suboptimal Viral Suppression Rates among HIV-Infected Children in Low-And Middle-Income Countries: A Meta-analysis,” Clinical Infectious Diseases, vol. 63, no. 12, 2016, doi: 10.1093/cid/ciw645.

[13] M. Braun et al., “Inadequate coordination of maternal and infant HIV services detrimentally affects early infant diagnosis outcomes in lilongwe, Malawi,” J Acquir Immune Defic Syndr (1988), vol. 56, no. 5, 2011, doi: 10.1097/QAI.0b013e31820a7f2f.

[14] C. A. Rodriguez et al., “Lifetime Changes in CD4 T-cell count, Viral Load Suppression and Adherence among Adolescents Living with HIV in Urban Peru,” Pediatric Infectious Disease Journal, vol. 39, no. 1, 2020, doi: 10.1097/INF.0000000000002501.

[15] O. A. Adejumo, K. M. Malee, P. Ryscavage, S. J. Hunter, and B. O. Taiwo, “Contemporary issues on the epidemiology and antiretroviral adherence of HIV-infected adolescents in sub-Saharan Africa: A narrative review,” J Int AIDS Soc, vol. 18, no. 1, 2015, doi: 10.7448/IAS.18.1.20049.

[16] S. H. Kim, S. M. Gerver, S. Fidler, and H. Ward, “Adherence to antiretroviral therapy in adolescents living with HIV: Systematic review and meta-analysis,” AIDS, vol. 28, no. 13, 2014, doi: 10.1097/QAD.0000000000000316.

[17] World Health Organization, “Implementing WHO evidence-based interventions for adolescents and young adults living with and affected by HIV,” Geneva, 2024.

[18] C. Oduenyi, E. Ugwa, Z. Ojukwu, and J. Ojukwu-Ajasigwe, “An Exploratory Study of Stigma and Discrimination among People Living with HIV/ AIDS in Abuja Municipal Area Council, Nigeria.,” Afr J Reprod Health, vol. 23, no. 1, pp. 88–99, Mar. 2019, doi: 10.29063/ajrh2019/v23i1.9.

[19] A. Cloete, L. C. Simbayi, S. C. Kalichman, A. Strebel, and N. Henda, “Stigma and discrimination experiences of HIV-positive men who have sex with men in Cape Town, South Africa,” AIDS Care, vol. 20, no. 9, pp. 1105–1110, Oct. 2008, doi: 10.1080/09540120701842720.

[20] E. I. Obeagu and G. U. Obeagu, “Unmasking the Truth: Addressing Stigma in the Fight Against HIV,” Elite Journal of Public Health, vol. 2, no. 1, pp. 8–22, 2024.

[21] B. X. Tran et al., “Understanding Global HIV Stigma and Discrimination: Are Contextual Factors Sufficiently Studied? (GAPRESEARCH).,” Int J Environ Res Public Health, vol. 16, no. 11, May 2019, doi: 10.3390/ijerph16111899.

[22] N. M. Else-Quest, A. M. French, and N. A. Telfer, “The intersectionality imperative: Calling in stigma and health research.,” Stigma Health, vol. 8, no. 3, pp. 269–278, Aug. 2023, doi: 10.1037/sah0000397.

[23] J. Y. Chow et al., “Peru’s HIV care continuum among men who have sex with men and transgender women: opportunities to optimize treatment and prevention.,” Int J STD AIDS, vol. 27, no. 12, pp. 1039–1048, Oct. 2016, doi: 10.1177/0956462416645727.

[24] A. Piñeirúa et al., “The HIV care continuum in Latin America: challenges and opportunities,” Lancet Infect Dis, vol. 15, no. 7, pp. 833–839, Jul. 2015, doi: 10.1016/S1473-3099(15)00108-5.

[25] Global Network of People Living with HIV, “Informe Multipaís índice de Estigma y Discriminación en Personas con VIH 2.0 Bolivia, Ecuador, Nicaragua, Perú y Paraguay,” 2023.

[26] K. Gabbidon, T. Chenneville, V. Earnshaw, and H. Drake, “Intersectional stigma and developmental competence among youth living with HIV,” J Fam Theory Rev, vol. 14, no. 3, pp. 520–536, Sep. 2022, doi: 10.1111/jftr.12468.

[27] J. R. de C. M. Ayres et al., “Vulnerability, human rights, and comprehensive health care needs of young people living with HIV/AIDS.,” Am J Public Health, vol. 96, no. 6, pp. 1001–6, Jun. 2006, doi: 10.2105/AJPH.2004.060905.

[28] A. L. Stangl et al., “The Health Stigma and Discrimination Framework: a global, crosscutting framework to inform research, intervention development, and policy on health-related stigmas.,” BMC Med, vol. 17, no. 1, p. 31, Feb. 2019, doi: 10.1186/s12916-019-1271-3.

[29] UNAIDS, “Country Fact Sheet: Peru,” 2023. Accessed: Oct. 23, 2024. [Online]. Available: https://www.unaids.org/en/regionscountries/countries/peru

[30] M. E. Perla et al., “Genital Tract Infections, Bacterial Vaginosis, HIV, and Reproductive Health Issues among Lima-Based Clandestine Female Sex Workers,” Infect Dis Obstet Gynecol, vol. 2012, pp. 1–9, 2012, doi: 10.1155/2012/739624.

[31] Ministerio del Perú, “Situación epidemiológica del VIH-Sida en el Perú,” 2021.

[32] S. Cousins, “A complex epidemic prevents Peru reaching HIV goals.,” Lancet HIV, vol. 6, no. 11, pp. e733– e734, Nov. 2019, doi: 10.1016/S2352-3018(19)30351-0.

[33] IOM Peru, “Encuesta Bioconductual (BBS) en Migrantes Venezolanos que viven en Lima/Callao y Trujillo,” Lima, 2023. Accessed: Feb. 16, 2025. [Online]. Available: https://www.r4v.info/es/document/reporte-final-encuestas-bioconductual-bbs-en-migrantes-venezolanos-que-viven-en-limacallao

[34] A. Tong, P. Sainsbury, and J. Craig, “Consolidated criteria for reporting qualitative research (COREQ): a 32-item checklist for interviews and focus groups,” International Journal for Quality in Health Care, vol. 19, no. 6, pp. 349–357, Sep. 2007, doi: 10.1093/intqhc/mzm042.

[35] “Dedoose Version 9.0; web application for managing; analyzing; and presenting qualitative and mixed method research data,” 2023, SocioCultural Research Consultants, LLC, Los Angeles, CA.

[36] L. J. Goldsmith, “Using framework analysis in applied qualitative research,” Qualitative Report, vol. 26, no. 6, 2021, doi: 10.46743/2160-3715/2021.5011.

[37] S. Klingberg, R. E. Stalmeijer, and L. Varpio, “Using framework analysis methods for qualitative research: AMEE Guide No. 164,” Med Teach, vol. 46, no. 5, 2024, doi: 10.1080/0142159X.2023.2259073.

[38] B. G. Link and J. Phelan, “Stigma power.,” Soc Sci Med, vol. 103, pp. 24–32, Feb. 2014, doi: 10.1016/j.socscimed.2013.07.035.

[39] J. Lucas, H.-Y. Ho, and K. Kerns, “Power, Status, and Stigma: Their Implications for Health,” in The Oxford Handbook of Stigma, Discrimination, and Health, Oxford University Press, 2017, pp. 69–84. doi: 10.1093/oxfordhb/9780190243470.013.15.

[40] T. Chenneville, “Formative HIV Research With Youth in Kenya: Findings From a Psychosocial Needs Assessment,” Journal of the Association of Nurses in AIDS Care, vol. 28, no. 3, pp. 443–449, May 2017, doi: 10.1016/j.jana.2016.11.004.

[41] M. L. Ekstrand, J. Ramakrishna, S. Bharat, and E. Heylen, “Prevalence and drivers of HIV stigma among health providers in urban India: implications for interventions.,” J Int AIDS Soc, vol. 16, no. 3 Suppl 2, p. 18717, Nov. 2013, doi: 10.7448/IAS.16.3.18717.

[42] J. Tattsbridge, C. Wiskin, G. De Wildt, A. Clavé Llavall and C. Ramal-Asayag, “HIV understanding, experiences and perceptions of HIV-positive men who have sex with men in Amazonian Peru: A qualitative study,” BMC Public Health, vol. 20, no. 1, 2020, doi: 10.1186/s12889-020-08745-y.

[43] D. Valencia-Garcia, D. Rao, L. Strick, and J. M. Simoni, “Women’s experiences with HIV-related stigma from health care providers in Lima, Peru: ‘I would rather die than go back for care,’” Health Care Women Int, vol. 38, no. 2, 2017, doi: 10.1080/07399332.2016.1217863.

[44] N. K. Fauk, P. R. Ward, K. Hawke, and L. Mwanri, “HIV Stigma and Discrimination: Perspectives and Personal Experiences of Healthcare Providers in Yogyakarta and Belu, Indonesia.,” Front Med (Lausanne), vol. 8, p. 625787, 2021, doi: 10.3389/fmed.2021.625787.

[45] P. Bravo, A. Edwards, S. Rollnick, and G. Elwyn, “Tough decisions faced by people living with HIV: a literature review of psychosocial problems.,” AIDS Rev, vol. 12, no. 2, pp. 76–88, 2010.

[46] B. Fatoki, “Understanding the Causes and Effects of Stigma and Discrimination in the Lives of HIV People Living with HIV/AIDS: Qualitative Study,” J AIDS Clin Res, vol. 7, no. 12, 2016, doi: 10.4172/2155-6113.1000635.

[47] S. Rueda et al., “Examining the associations between HIV-related stigma and health outcomes in people living with HIV/AIDS: a series of meta-analyses,” BMJ Open, vol. 6, no. 7, 2016, doi: 10.1136/BMJOPEN-2016-011453.

[48] K. Gabbidon, T. Chenneville, V. Earnshaw, and H. Drake, “Intersectional stigma and developmental competence among youth living with HIV,” J Fam Theory Rev, 2022, doi: 10.1111/jftr.12468.

[49] D. Wight, E. Wimbush, R. Jepson, and L. Doi, “Six steps in quality intervention development (6SQuID),” J Epidemiol Community Health (1978), vol. 70, no. 5, pp. 520–525, May 2016, doi: 10.1136/jech-2015-205952.

[50] T. Chenneville, K. Kosyluk, K. Gabbidon, M. Franke, D. Serpas, and J. T. Galea, “Positive, Open, Proud: an adapted disclosure-based intervention to reduce HIV stigma.,” Front Glob Womens Health, vol. 5, p. 1469465, 2024, doi: 10.3389/fgwh.2024.1469465.

[51] N. Rüsch and M. Kösters, “Honest, Open, Proud to support disclosure decisions and to decrease stigma’s impact among people with mental illness: conceptual review and meta-analysis of program efficacy.,” Soc Psychiatry Psychiatr Epidemiol, vol. 56, no. 9, pp. 1513–1526, Sep. 2021, doi: 10.1007/s00127-021-02076-y.

[52] G. Z. Andersson et al., “Stigma reduction interventions in people living with HIV to improve health-related quality of life.,” Lancet HIV, vol. 7, no. 2, pp. e129–e140, Feb. 2020, doi: 10.1016/S2352-3018(19)30343-1.

[53] P. Bravo, A. Edwards, S. Rollnick, and G. Elwyn, “Tough decisions faced by people living with HIV: A literature review of psychosocial problems,” 2010.

[54] B. Fatoki, “Understanding the Causes and Effects of Stigma and Discrimination in the Lives of HIV People Living with HIV/AIDS: Qualitative Study,” J AIDS Clin Res, vol. 7, no. 12, 2016, doi: 10.4172/2155-6113.1000635.

