## Supplemental Table 1 for "Drivers, Facilitators, and Impacts of Stigma Among Youth Living with HIV in Lima, Peru"

**Supplemental Table 1. Exemplary quotes: drivers, perpetuators, and outcomes of HIV-related stigma**

| | |
|---|---|
| **Drivers of stigma** | |
| Lack of information about HIV | *I think the lack of information leads people to be afraid, afraid of getting close to [someone with HIV]. Even though it's 2023, there's still a real lack of information. (Healthcare provider)* |
| | *It's because someone's ignorant, they don't know that HIV has, not a cure, but treatment that allows you to live much longer, that you can be a normal person, you can even have a family. (YPLWH, TGW)* |
| Misinformation about HIV | *Misinformation isn't just ignorance, but also erroneous beliefs shared within a circle. (YPLWH, GBMSM)* |
| **Key power groups perpetuating stigma** | |
| Parents & older generations | *I think [perpetuators are] the older people, because they lived during a time that's very different from what we young people are experiencing today. They were alive at a time when HIV was elevated and quality of life was low and treatment and diagnosis were very different from today. (YPLWH, GBMSM)* |
| | *From my perspective it's more generational. I've had young doctors and older doctors and at least I do feel there's a difference in the information, in their ways, their treatment, their use of terminology, even their willingness, empathy towards people. At least from my experience. I think it's a generational thing because of culture. (HIV advocate)* |
| Educators | *At least when you start secondary school, these are topics that are discussed and it's the information they always give you, it's transmitted like this, etc. etc. But they don't teach you the human aspect, like if John has this condition it's possible he can have a normal life, he can have a family, he can work, there's no type of impediments. (YPLWH, cisgender woman)* |
| | *I think there are many educators who when they talk about HIV in class, they continue to do so presenting HIV as it was many years ago. They don't inform as they should. (Healthcare provider)* |
| | *Also at school I remember that one day the teacher started talking about HIV and about how people who have HIV can't do everything, that they have to stay out of society. (YPLWH, GBMSM)* |
| | *I think that people who have a job where they're in charge of many people, for example teachers … they do have to be careful. For example I've had professors at university who've talked openly about HIV. I don't know why but they've given completely wrong opinions, with information from a long time ago. And I don't know if there's a law or something but you have to be very careful with that, the issue of the opinions of people who have a large audience. Because for example with professors, what they say, if they say A, the students think that's 100% true and they believe it. And that's bad. (YPLWH, GBMSM)* |
| Healthcare providers | *As a doctor, what I see from my position, there's not a friendly environment. For example, the Infectious Disease department is different, but in dental areas, gynecology, [PLWH] are seen last so they don't contaminate the office. They're attended to very poorly… treated as if they're terminally ill so it's not worth investing effort, money, and supplies in them. (Healthcare provider)* |
| | *I always heard those comments, why should [PLWH] get treatment if it's their fault, they're careless, they got it because they were irresponsible. And I made the comment, 'wow say that to a diabetic right.' I mean how irresponsible. And my mom, a doctor, participated in this type of commentary. (YPLWH, GBMSM)* |
| | *It's not so much because of bad information, not knowing, many of us have the knowledge. But even so there's still those types of comments, actions. And if they're coming from someone who apparently knows about health topics, people listening are going to say 'yeah, he's right, if he says it he's right.' (Healthcare provider)* |
| Other key power groups | *The ultraconservatives like the evangelicals, the protestants, Mormons, the Catholics not as much, but the Mormons, evangelicals. They're the ones who believe that being gay is a sin but if it's with HIV even worse. I mean you have to disappear, to the point of aggression. (YPLWH, GBMSM)* |
| | *If they find out at work that a person has HIV, they don't move up anymore, they get no more promotions. (Infectious Disease Specialist)* |
| **Outcomes of stigma** | |
| Fear and anxiety | *My fear is at work, since the [boss] has these stereotypes, I don't want her to find out about my diagnosis. As I was saying, I go to the doctor a lot, to my appointments, my check-ups. My colleagues know that I go to the doctor but they don't know why. But since people aren't stupid, I don't want them to out me and for the boss to ask for a test, because for a while she's been saying she's going to ask for a test. So I turn every color in the book, I start sweating, the nerves… (YPLWH, GBMSM, Venezuelan migrant)* |
| Shame, guilt, and loneliness | *Also the issue of shame… the fact that your partner could feel, you feel it without them saying anything, you feel a change in them, after you tell them. Maybe before you enjoyed sex without any worries, it was sex, without having anything there to stop you. And once you tell them, there might be a change in their attitude towards sex and you feel it. The worst is that they don't say anything but you feel it. So that's what feeds the sensation of shame I think, that even though the information about undetectable is* |

| | |
|---|---|
| | *untransmittable exists, people still don't get it. There's always that suspicion of what would happen, because it's not 100%, that 1% maybe. (HIV advocate)* |
| | *To be honest, it's what happens to me everyday. When I told my sister, who I love with all my heart, the first thing she did was separate me from my nieces, she isolated me, she didn't want me to get close or anything. I don't live with them, and when I started to tell my mom little by little, she started to become distant. So since then I've started to feel uncomfortable and I started to feel bad about myself, I felt disgusted with myself. I've had to hide everything that I use, everything that I do, I feel like a fish out of water. I have to hide everything that's mine or whatever I touch and pretend that nothing's wrong. I even have my schedule for taking my medicine and I try not to drink or smoke because I feel like it would just make it worse. And people my age don't get it, 'why aren't you drinking, why do you have, what do you have, what's wrong?' and I say, 'no it's nothing, it's just for my blood pressure, my diabetes, it's nothing.' So that's happened a lot, both at health centers, in my family, and it's really really awful. (YPLWH, FSW)* |
| | *Finding out my diagnosis, because I've had a lot of experience with people who have the same diagnosis as I do now, because of my career path, I mean I was seeing patients. And for me it was normal to read in the chart that a patient was seropositive. I didn't think twice about it. But now that's me, I don't understand why I feel like this, why this lack of confidence, this lack of self-esteem. I don't know why this diagnosis hits so hard. (YPLWH, GBMSM, Venezuelan migrant)* |
| Health | *I was being seen at a clinic where I was first diagnosed and the doctor told me not to come back, that he doesn't see patients like me, that [HIV] is only dealt with at hospitals. It was really shocking for me, and worse because I suffer from depression, so everything came pouring over me. (YPLWH, cisgender woman)* |
| | *Me, truthfully, I tried to kill myself after my diagnosis. I tried to kill myself twice, more than anything because, I was ok until the father of my child took him away from me. He said, 'you think I'm going to let someone with HIV be with my child?' And I said, 'how mean.' He let him be inside of me, I've made sure he was healthy and now he's taking him from me. He took him from me when he was one year old… I ingested poison. Truthfully I didn't think about my other child, about anything, I wanted to kill myself. For those four years I didn't take my medicine, I thought, 'what do I have to live for?' (YPLWH, FSW)* |
| | *People lose self-esteem, they say you know what, why should I take [my medicine], why should I keep living. And that's when you start losing weight, get depressed and that can lead to death. (HIV advocate)* |
| | *A lot of times there's also an emotional impact, emotional instability, mental health disorders, that end up having an impact on treatment adherence, it leads to a lot of abandoning of treatment. The support that a patient has in their environment is very important, because it can have a positive impact, or at the other extreme a very negative impact. (Healthcare provider)* |
| Social | *[Stigma] is one of the reasons I started my own business. I said, 'wow, I'm going to have to go through this and it's what I least want and I don't know how to deal with it.' So it was better to just, since my mom has her own business, I started to invest with her and now I have my own business where I can choose to be there or not. (YPLWH, GBMSM)* |
| | *In my case for example, I had depression, I missed classes, the medicine made me sleepy, I was tired all the time. And the professor felt I wasn't doing my best, he thought I was lazy because I was missing classes. (HIV advocate)* |
| | *In some patients [the effects of HIV-stigma] could be thinking and feeling that your life isn't going to be the same, in relationships for example, thinking that you're going to be alone and you're not going to be able to have a family if you're homosexual… that your life is going to be limited in terms of partners. I've seen that a lot. (Healthcare provider)* |
